# KIM-1/TIM-1 is a Receptor for SARS-CoV-2 in Lung and Kidney

**DOI:** 10.1101/2020.09.16.20190694

**Authors:** Yutaro Mori, Corby Fink, Takaharu Ichimura, Keisuke Sako, Makiko Mori, Nathan N. Lee, Philipp Aschauer, Krishna M. Padmanabha Das, SoonGweon Hong, Minsun Song, Robert F. Padera, Astrid Weins, Luke P. Lee, Mahmoud L. Nasr, Gregory A. Dekaban, Jimmy D. Dikeakos, Joseph V. Bonventre

## Abstract

SARS-CoV-2 precipitates respiratory distress by infection of airway epithelial cells and is often accompanied by acute kidney injury. We report that Kidney Injury Molecule-1/T cell immunoglobulin mucin domain 1 (KIM-1/TIM-1) is expressed in lung and kidney epithelial cells in COVID-19 patients and is a receptor for SARS-CoV-2. Human and mouse lung and kidney epithelial cells express KIM-1 and endocytose nanoparticles displaying the SARS-CoV-2 spike protein (virosomes). Uptake was inhibited by anti-KIM-1 antibodies and TW-37, a newly discovered inhibitor of KIM-1-mediated endocytosis. Enhanced KIM-1 expression by human kidney tubuloids increased uptake of virosomes. KIM-1 binds to the SARS-CoV-2 Spike protein *in vitro*. KIM-1 expressing cells, not expressing angiotensin-converting enzyme 2 (ACE2), are permissive to SARS-CoV-2 infection. Thus, KIM-1 is an alternative receptor to ACE2 for SARS-CoV-2. KIM-1 targeted therapeutics may prevent and/or treat COVID-19.

## Introduction

Coronavirus disease 2019 (COVID-19) caused by SARS-CoV-2 ^1^ was first reported at Wuhan in China in 2019 ^2^. The disease has since reached pandemic proportions ^3,4^. SARS-CoV-2-related respiratory failure and acute kidney injury (AKI) are significant complications of infection ^5-7^ and are associated with high morbidity and mortality ^8,9^. **K**idney **I**njury **M**olecule**-1** (KIM-1), also identified as Hepatitis A Virus Cellular Receptor 1 (HAVCR1) in hepatocytes ^10^, and T-cell immunoglobulin and mucin domain 1 (TIM-1), was identified by our group as the most upregulated protein in the kidney proximal tubule after a wide variety of injurious influences including ischemia, nephrotoxicants, sepsis, and immune-related injury and its cleaved ectodomain is often used as a blood and urine marker for kidney injury ^11-16^. KIM-1 has been reported as a receptor for Ebola virus ^17^ and Dengue virus ^18^. KIM-1 also facilitates cellular uptake of West Nile virus ^19^. KIM-1 can mediate internalization and transduction of Marburg virus glycoprotein (GP) and full-length Ebola virus GP pseudovirions into human mucosal epithelia from the trachea ^17^. KIM-1-mediated infection was efficiently inhibited by anti-human KIM-1 IgV domain-specific monoclonal antibody ARD5 ^20^. In the kidney, KIM-1 acts as a receptor for phosphatidylserine exposed on the surface of apoptotic cells and for oxidized lipids ^21^. Once ligands bind to KIM-1, they are internalized by phagocytosis or endocytosis. We have recently identified TW-37 as an inhibitor of KIM-1-mediated oxidized lipid and palmitic acid (bound to albumin) uptake ^22^.

The SARS-CoV-2 envelope contains a spike (S) glycoprotein, consisting of S1 and S2 subunits ^23^. After infection, the trimeric S protein is cleaved into the two subunits, and S1, which contains the receptor-binding domain, binds to angiotensin-converting enzyme 2 (ACE2) and is internalized by lung epithelium. In addition to ACE2, carcinoembryonic antigen-related cell adhesion molecule (CEACAM) is known to be a receptor for SARS-CoV ^24^. KIM-1 has an N-terminus conserved IgV domain with high homology with CEACAM’s IgV domain to which the S1 subunit binds ^25^. T-cell immunoglobulin and mucin domain 3 (TIM-3), another KIM/TIM-family member protein, forms a heterodimer with CEACAM ^26^. Although KIM-1 is expressed at much higher levels in the kidney, it has also been reported to be expressed in human primary airway epithelial cells in pulmonary disease, human non-small-cell lung cancer, and human lung adenocarcinoma A549 cells ^27,28^. KIM-1 is also expressed in Vero E6 monkey kidney cells used for *in vitro* expansion and maintenance of Coronaviruses, including SARS-CoV-2 and other viruses^29^.

Here, we show that KIM-1 acts as a receptor for SARS-CoV-2 both in lung and kidney epithelia. KIM-1 expression in alveolar epithelium co-localizes in cells with SARS-CoV-2 nucleocapsid protein in post-mortem lung samples from COVID-19 patients. KIM-1 can bind to and mediate the internalization of liposomal nanoparticles displaying the SARS-CoV-2 spike protein ectodomain on their surface (virosomes) *in vitro*. The internalization of virosomes is inhibited both by anti-KIM-1 antibodies and TW-37. Molecular binding between KIM-1 and SARS-CoV-2 Spike protein was detected. KIM-1 expressing cells that did not express angiotensin-converting enzyme 2 (ACE2) were infected by SARS-CoV-2. Thus, KIM-1 may play an essential role in viral infection and can be a potential therapeutic target to mitigate the effects of SARS-CoV-2 in both lung and kidney.

## RESULTS

### KIM-1 is expressed in COVID-19 patient autopsy lung samples

KIM-1 was expressed in the post-mortem lungs of 10 of 11 patients who died following SARS-CoV-2 infection. KIM-1 was present in pan-cytokeratin-positive alveolar epithelial cells that were dislodged and formed debris-like clusters in most cases. In two patients (Patient 1 and 2), positive staining for SARS-CoV-2 nucleocapsid protein was present in KIM-1-positive and pan-cytokeratin-positive alveolar epithelial cells (**Fig. 1a**). KIM-1 (detected with two antibodies (AF1750 and AKG7)) was also co-localized with surfactant protein C (**Fig. 1a**). KIM-1 was detected using two different antibodies (AF1750 polyclonal and AKG7 monoclonal ^20^). ACE2, a well-known receptor for SARS-CoV-2, was expressed and, in patient 1, is seen co-localized with KIM-1 and pan-cytokeratin (**Fig. 1a**). KIM-1 expression was very weak in non-COVID-19 patient lung autopsy samples (**Supplementary Fig. 1**).

**Figure 1.**
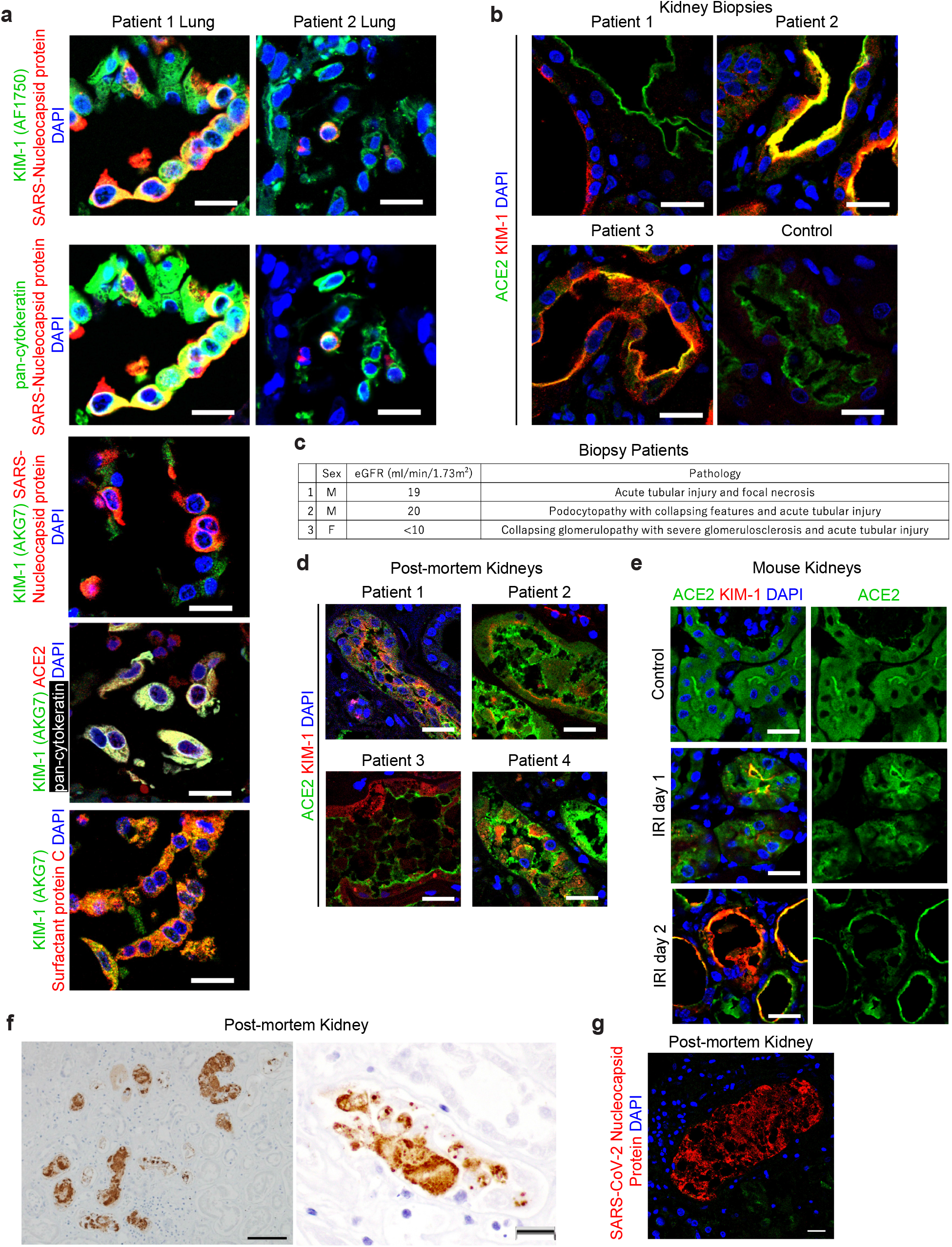
KIM-1 is expressed in COVID-19 patient alveolar and kidney epithelia. **(a)** Representative immunostaining of KIM-1, SARS-CoV-2 nucleocapsid protein and pan-cytokeratin (top two rows), immunostaining of KIM-1 and SARS-CoV-2 nucleocapsid protein (3rd row), and immunostaining of ACE2, KIM-1, pan-cytokeratin and surfactant protein (bottom 2 panels) in kidney biopsy samples from COVID-19 patients. DAPI staining marks the nuclei in this and other panels. Scale bars, 20 μm. **(b)** Immunostaining of KIM-1 and ACE2 in a post-mortem lung sample from COVID-19 patients. Scale bar, 20 μm. **(c)** Patient information for three COVID-19-associated AKI kidney biopsy samples. The patients’ ages were at a range of 45-60 years old. **(d)** Immunostaining of KIM-1 and ACE2 in representative post-mortem kidney biopsy samples from four COVID-19 patients. Scale bars, 20 μm. **(e)** Immunostaining of KIM-1 and ACE2 post ischemia-reperfusion injury (IRI) in mouse kidneys. Scale bars, 20 μm. **(f)** Immunohistochemistry staining of SARS-CoV-2 nucleocapsid protein in a COVID-19-associated AKI patient. Scale bars, 50 μm (left panel), 20 μm (right panel). **(g)** Immunostaining of SARS-CoV-2 nucleocapsid protein in a COVID-19 post-mortem kidney section from the patient where immunohistochemisty is shown in **Fig 1f**. Scale bar, 20 μm.

### KIM-1 is expressed in COVID-19 patient biopsy and autopsy kidney proximal tubules

Since AKI is a frequent manifestation of COVID-19, we evaluated KIM-1 expression in COVID-19 patient kidney biopsies from 3 live patients with AKI who survived their hospitalization and 30 post-mortem kidney samples from patients with documented SARS-CoV-2 infection during their terminal hospitalization. KIM-1 expression was seen in all 3 biopsy cases, with approximately 10% of proximal tubules staining positive in both the cortex and outer medulla (**Fig. 1b and 1c**). Estimated glomerular filtration rate (GFR) and brief descriptions of the predominant renal pathology at the time of biopsy are presented in **Fig. 1c**. KIM-1 positive tubules were dilated with some tubules containing cellular debris. ACE2 is widely expressed on the apical side of proximal tubules in the same regions as KIM-1. KIM-1 positive tubules tended to have reduced ACE2 staining, suggesting that ACE2 might be down-regulated as tubules dedifferentiate and enhance KIM-1 expression in **Fig. 1b**. We detected KIM-1 in 14 of 30 autopsy cases (**Fig. 1d**). In some patients, KIM-1 was expressed in relatively well-preserved tubules; however, in most kidneys with KIM-1 expression, the protein was localized in exfoliated tubular cells due to postmortem artifacts. In some cases, ACE2 was also detected in those exfoliated cells. Thus, in COVID-19 patient kidneys, KIM-1 was frequently expressed, and significant tubular injury and KIM-1 expression were associated with less ACE2 expression in the proximal tubules. We compared this reciprocal response between KIM-1 and ACE2 to a well-established mouse model of AKI. When KIM-1 and ACE2 were stained in post-ischemia-reperfusion injury (IRI) in mouse kidneys, ACE2 was down-regulated when KIM-1 was increased in flattened epithelial cells of the proximal tubules (**Fig. 1e**). In one of the autopsy kidneys from a male in his fifties with an eGFR of 16 ml/min/1.73m^2^, SARS-CoV-2 nucleocapsid protein was observed in focal areas of proximal tubules and intraluminal casts of cellular debris (**Fig. 1f, g**).

### The role of KIM-1 in promoting SARS-CoV-2 entry in lung alveolar epithelial cells

SARS-CoV-2 virosomes were constructed of phospholipid liposomes conjugated to His-tagged spike ectodomain on their surfaces (**Fig. 2a**), and uptake of these fluorescently (Dil) labeled biomimetic viruses (virosomes, **Fig. 2a**) was assessed *in vitro* after confirming virosomes and control empty liposomes were equally labeled by DiI (**Supplementary Fig. 2**). Virosomes do not contain phosphatidylserine or phosphatidylethanolamine that are established ligands for KIM-1 ^21^. A549 cells, adenocarcinoma human alveolar basal epithelial cells, express KIM-1 and endocytosed SARS-CoV-2 virosomes while there was minimal uptake of unconjugated labelled liposomes (**Fig. 2b**) as assessed by confocal microscopy and quantified by flow cytometry. Cellular uptake of virosomes was efficiently inhibited by treatment with anti-KIM-1 IgG or with TW-37, our newly discovered inhibitor for KIM-1-mediated endocytosis ^22^ (**Fig. 2c**). Virosomes were taken up by 92.3% ± 1.1% of A549 cells, and anti-KIM-1 antibodies and TW-37 decreased the uptake to 44.9% ± 17.0% and 3.5% ± 0.2%, respectively quantitated by flow cytometry (**Fig. 2d**). ACE2 was also expressed in A549 cells and was not altered in expression by anti-KIM-1 antibody (**Fig. 2e**). We also analyzed the uptake of virosomes by mouse primary lung epithelial cells. Cells were isolated from wild-type mice and mice carrying a mutation in the KIM-1 mucin domain (KIM-1^Δmucin^ mice) ^30^. KIM-1^Δmucin^ serves as a functional knockout with reduced epithelial cell uptake of known KIM-1 ligands ^16,31^. Wild-type mouse primary lung epithelial cells took virosomes up efficiently, similar to A549 cells. Treatment with anti-KIM-1 antibody or TW-37 (10 µM) dramatically decreased the uptake of virosomes, and KIM-1^Δmucin^ cells showed significantly less uptake than wild-type cells (**Fig. 2f, g**).

**Figure 2.**
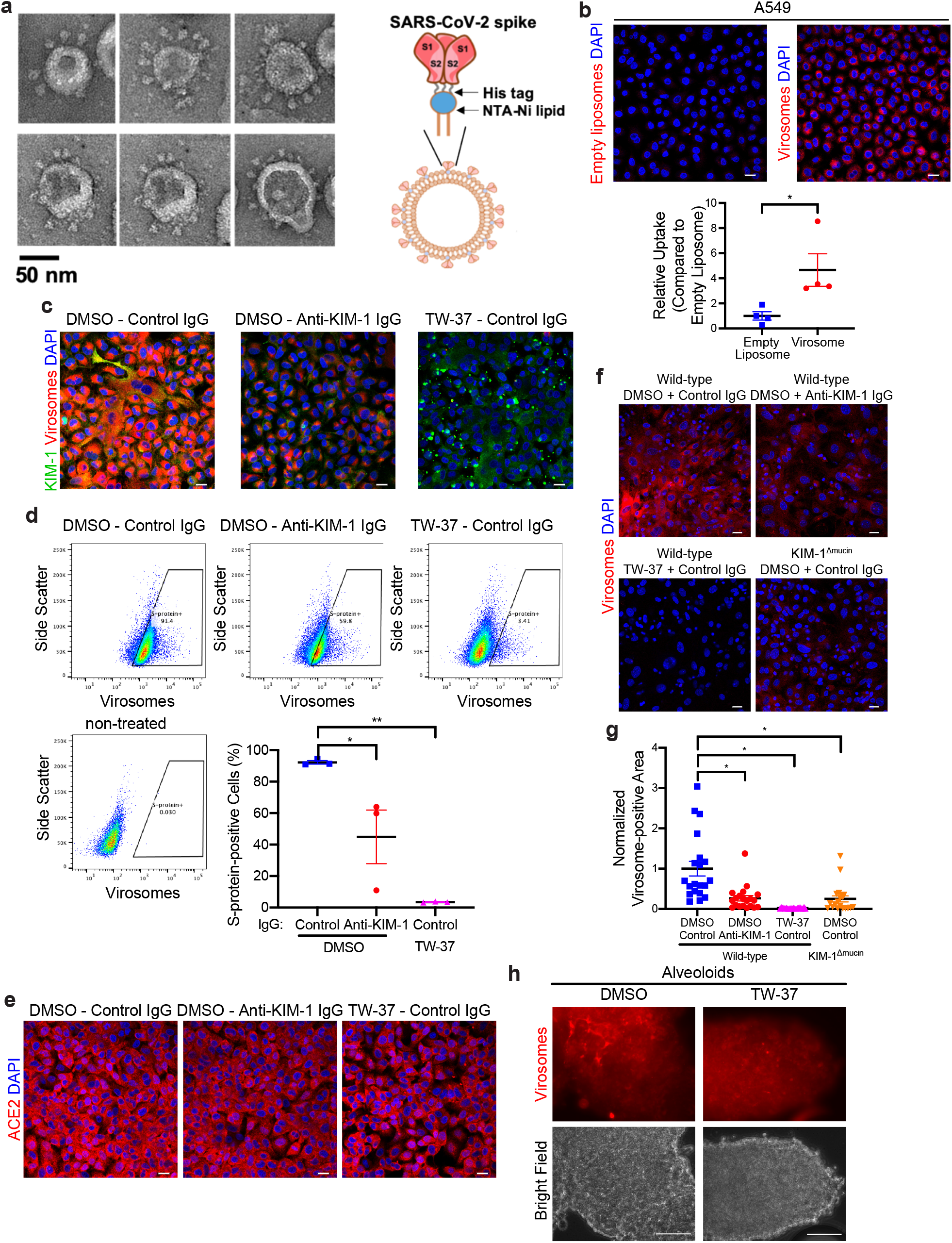
SARS-CoV-2 virosomes containing the spike protein with both S1 and S2 subunits are internalized by lung epithelial cells and A549 adenocarcinoma human alveolar basal epithelial cells in a KIM-1-dependent manner. **(a)** Negative-stain electron micrograph of SARS-CoV-2 virosomes displaying the spike ectodomain. The spike His-tag was bind to the nickel-nitrilotriacetic acid (Ni-NTA) lipids. **(b)** Top: A549 cell internalization of DiI-labeled virosomes as compared to control empty liposomes with equivalent Dil (0.5 nM for each). Scale bars, 20 μm. Bottom: Quantification of internalized DiI-positive cells by flow cytometry. *p=0.017. **(c)** Internalization of DiI-labeled virosomes by A549 cells in the presence of control mouse IgG, anti-KIM-1 (3F4 and AKG7) or TW-37, a specific inhibitor for KIM-1, or control DMSO, immunostained with KIM-1 antibody (green). Scale bars, 20 μm. **(d)** Quantification of internalized DiI-labeled virosomes by A549 cells as measured by flow cytometry. ^*^p=0.0334, ^**^p=0.0017. **(e)** ACE2 immunostaining after uptake of DiI-labeled virosomes into A549 cells. Scale bars, 20 μm. **(f)** Internalization of DiI-labeled virosomes by mouse primary lung epithelial cells from wild-type mice or KIM-1^Δmucin^ mice after pretreatment with control IgG, anti-KIM-1 IgG (AF1817) or TW-37 or control DMSO. Scale bars, 20 μm. **(g)** Quantification of internalized DiI-labeled virosomes by mouse primary lung epithelial cells treated as described in **Fig. 2f** and measured by ImageJ. Virosome-positive areas were normalized to the average of wild-type cells treated with DMSO and control IgG. Ten fields were analyzed in two independent experiments. ^*^p<0.0001. **(h)** Internalization of DiI-labeled virosomes by human alveoloids with or without TW-37. Scale bar: 100 μm.

To study further the KIM-1-mediated SARS-CoV-2 entry into lung epithelial cells, we developed a 3D model of human alveoli (“alveoloids”) using A549 cells (**Fig. 2h**). Fluorescently-labeled SARS-CoV-2 spike protein conjugated virosomes were taken up by alveoloids, and this uptake was inhibited by TW-37 (10 µM) treatment (**Fig. 2h**).

### The role of KIM-1 in the promotion of SARS-CoV-2 entry into kidney cells

A stably transfected LLC-PK1 pig kidney cell line expressing human KIM-1 (hKIM-1-LLC-PK1), and a control line without KIM-1 expression (pcDNA3-LLC-PK1), were incubated with fluorescently-labeled SARS-CoV-2 spike protein conjugated virosomes (**Fig. 3a**). The hKIM-1-LLC-PK1 cells endocytosed SARS-CoV-2 virosomes in a KIM-1-dependent manner, whereas there was no uptake of unconjugated empty liposomes (**Fig. 3a**) as assessed by confocal microscopy. TW-37 (10 µM) pre-treatment for 30 min markedly reduced uptake of SARS-CoV-2 virosomes into hKIM-1-LLC-PK1 cells (**Fig. 3b**). To confirm virosome internalization in addition to cell surface attachment, we analyzed Z-stack confocal images of DiI-labeled virosome uptake by LLC-PK1 cells expressing human KIM-1 treated with or without TW-37 (**Fig. 3c**). TW-37 treatment decreased this internalization. Additionally, we tested virosome uptake by LLC-PK1 cells in a microfluidic channel with shear stress recapitulating the flow of formative urine. LLC-PK1 cells took up SARS-CoV-2 virosomes in a KIM-1-dependent manner as well, which was dramatically inhibited by TW-37 (**Fig. 3d**). Furthermore, we tested attachment and entry of GFP-labeled coronavirus-based SARS-CoV-2 pseudovirus (CoV2-01, SARS-CoV-2-S(GFP)) having SARS-CoV-2 Spike on their surface on LLC-PK1 cells stably expressing KIM-1 or control pcDNA in a co-cultured condition. Infection of this pseudovirus visualized by GFP was dominantly observed in hKIM-1-LLC-PK1 cells but not in pcDNA-LLC-PK1 cells (**Fig. 3e**). Additionally, soluble KIM-1 cleaved from hKIM-1-LLC-PK1 cells does not enhance infection on pcDNA-LLC-PK1 cells (**Fig. 3e**).

**Figure 3.**
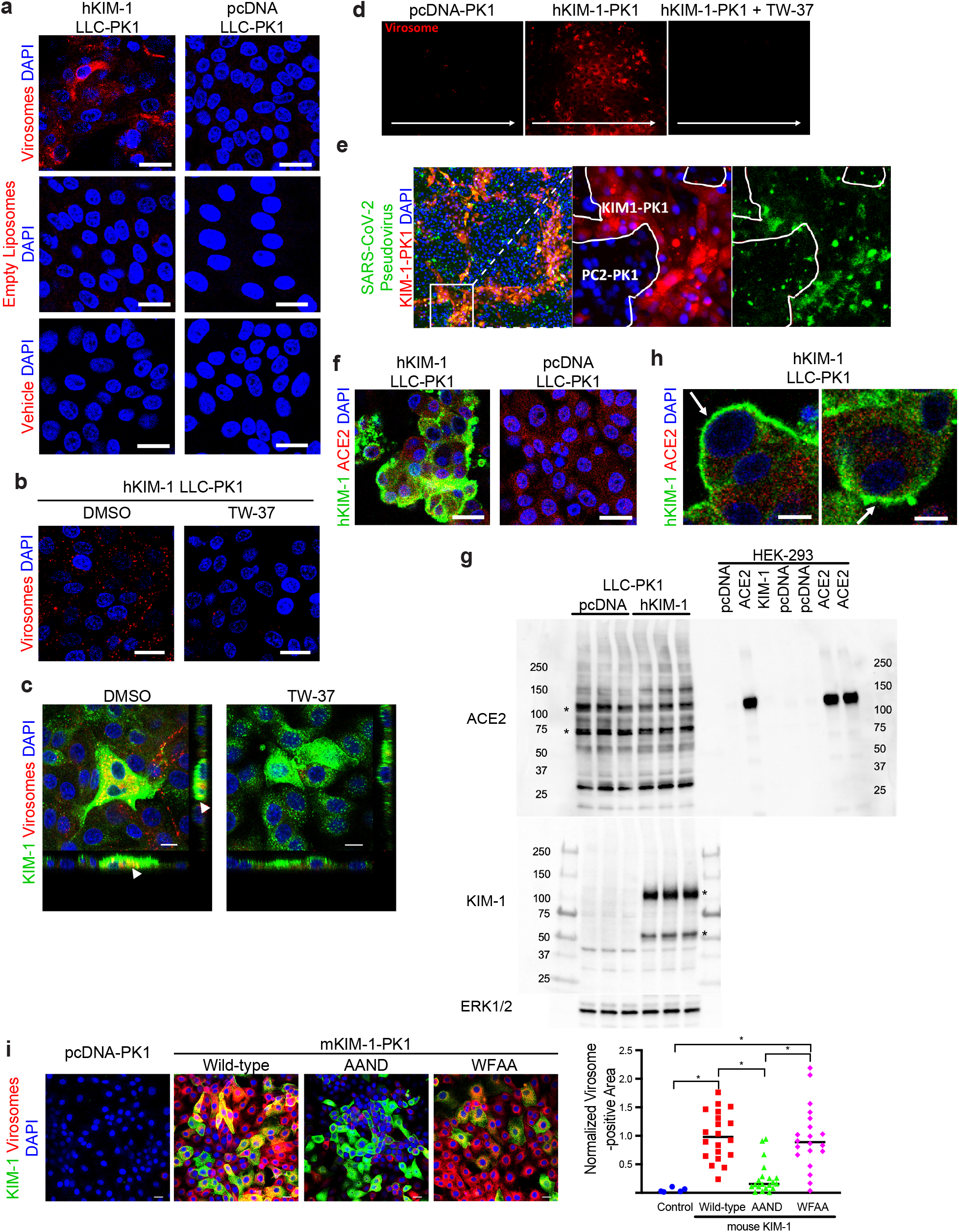
SARS-CoV-2 virosomes and pseudovirus are internalized by LLC-PK1 renal epithelial cells in a KIM-1-dependent manner. **(a)** Internalization of DiI-labeled virosomes or control Dil-labeled empty liposomes by LLC-PK1 cells stably expressing KIM-1 or control pcDNA. Scale bars, 20 μm. **(b)** Internalization assay of DiI-labeled virosomes on LLC-PK1 cells stably expressing human KIM-1 pretreated with TW-37 or control DMSO. Scale bars, 20 μm. **(c)** Z-stack analysis of KIM-1-expressing LLC-PK1 cells treated with DiI-labeled virosomes with or without TW-37 by confocal microscopy. Arrowheads indicate the internalized DiI-labeled virosomes. Scale bars: 10 μm. **(d)** Internalization of DiI-labeled virosomes by LLC-PK1 cells stably expressing KIM-1 or control pcDNA in a microfluidic channel with constant flow (0.2 dyn/cm^2^). Arrows indicate direction of flow. **(e)** After staining with an orange-color fluorescent dye, KIM-1 expressing cells and control pcDNA cells were mixed, co-cultured together, and exposed to GFP-tagged SARS-CoV-2 pseudovirus (CoV2-01, SARS-CoV-2-S(GFP)) for 12 hours. Red and Green signals were imaged 1.5 days after infection. **(f)** Immunostaining of KIM-1 (AKG7) and ACE2 in LLC-PK1 cells stably expressing human KIM-1 or control pcDNA. Scale bars, 20 μm. **(g)** Western blotting of ACE2, KIM-1 (cytoplasmic domain, Ab #195) and ERK1/2 for confirmation of equal loading of lanes on using extracts of LLC-PK1 cells stably expressing human KIM-1 or control pcDNA. Asterisks indicate the bands for each protein. **(h)** Immunostaining of KIM-1 and ACE2 on LLC-PK1 cells stably expressing human KIM-1. Arrows indicate that KIM-1 expression is located in cell surface. Scale bars, 10 μm. **(i)** Left: Internalization assay of DiI-labeled virosomes on LLC-PK1 cells stably expressing wild-type or mutant mouse KIM-1 (wild-type, AAND mutant of WNFD binding motif in Ig domain (amino acid residues 112-115) or WFAA mutant) or control pcDNA. Scale bars: 20 μm. Right: Quantification of internalized DiI-labeled virosomes measured by ImageJ. ^*^p<0.0001.

Both hKIM-1-LLC-PK1 and pcDNA3-LLC-PK1 cells stained positively for ACE2; however, much of the ACE2 was intracellular. pcDNA3-LLC-PK1 control cells appeared to have more intense and defuse ACE2 staining, even though the cells did not demonstrate significant uptake of the virosomes compared to the hKIM-1-LLC-PK1 cells (**Fig. 3f**). ACE2 protein expression was confirmed by western blot analysis of the cell lysates from both cell lines, whereas KIM-1 was expressed only in the hKIM-1-LLC-PK1 cells (**Fig. 3g**). ACE2 bands are located at approximately 75 kDa and 120 kDa ^32^. ACE2 and KIM-1 were co-stained and analyzed in high magnification confocal microscope images, ACE2 was localized to the cytoplasm with small amounts on the cell surface, while KIM-1 expression was primarily on the cell surface (arrows) (**Fig. 3h**). We tested uptake by LLC-PK1 cells expressing KIM-1 mutants of the binding motif, a four-amino acid motif (Tryptophan-Phenylalanine-Asparagine-Aspartic acid, WFND, amino acid residues 112-115) in its extracellular Ig domain that serves to mediate phosphatidylserine (PS) binding to verify the KIM-1’s motifs important for uptake of Spike. Cells expressed mouse KIM-1 with Alanine-Alanine (AA) replacing either WF or ND ^16^. Virosome uptake by the WFAA mutant KIM-1-expressing cells was similar to wild-type KIM-1 (WFND) expressing cells (**Fig. 3i**). By contrast, there was significantly less virosome uptake by AAND expressing cells. Thus the WF motif is necessary for uptake of Spike. This is different from the binding of KIM-1 to phosphatidylserine on apoptotic cells where modification of either the WF or ND amino acids eliminates binding ^16^.

### Three-dimensional human renal epithelial tubuloids take up virosomes

We have developed a method to generate epithelial KIM-1 expressing “tubuloids” from kidney tissue derived from human subjects ^22^. Human primary epithelial cell cultures were established from the non-tumor kidney tissue removed from patients with kidney cancer. Those primary cells were cultured in suspension with growth factors and Matrigel in non-adherent dishes (see Methods). KIM-1 expressing cells of human tubuloids took up DiI-labeled virosomes, but not DiI-labeled empty liposomes (**Fig. 4a**). With the uptake of spike conjugated virosomes, the tubuloids maintained their tubule-like three-dimensional structures as seen in phase-contrast images (**Fig. 4b**). Since well-developed tubuloids with polar epithelium reduce expression of KIM-1, likely due to progressive differentiation of the tubular epithelium, we enhanced human KIM-1 expression by infecting tubuloids with an adenovirus expressing KIM-1 (Adenovirus-KIM-1) or the control vector expressing β-galactosidase (β-GAL) (Adenovirus-β-GAL). Both viral vectors contained a GFP expression cDNA for tracing viral infection and transgene expression in the infected cells. After 48 hours of infection, we added DiI-labeled virosomes to the tubuloids (**Fig. 4c**). Adenovirus-KIM-1 infected GFP-positive tubuloid cells showed high uptake of virosomes (**Fig. 4c**, left) and robust expression of KIM-1 protein (**Fig. 4c**, right). The KIM-1 infected tubuloids showed polarized expression of KIM-1 similar to proximal tubules *in vivo* and expressed ACE2 (**Fig. 4d**). In contrast to the Adenovirus-KIM-1 infected cells, control Adenovirus-β-GAL infected tubuloids showed little KIM-1 staining and little uptake of virosomes or oxidized LDL (data not shown). The uptake of virosomes by the tubuloids overexpressing KIM-1 was dose-dependent (**Fig. 4e**). Quantification of ACE2 expression by flow cytometry on tubuloids infected by Adenovirus-KIM-1 or control Adenovirus-β-GAL, after digestion into single cells, revealed a mean 29.8% decrease on single-cell ACE2 expression secondary to Adenovirus-KIM-1 infection (**Fig. 4f**). Adenovirus-KIM-1 infection decreased Lotus tetragonolobus lectin (LTL) expression when compared with control Adenovirus-β-GAL treatment, indicating that KIM-1 expression induces de-differentiation of human tubule epithelial cells in tubuloids (**Supplementary Fig. 3**). To demonstrate the effect of KIM-1 on virosome uptake also in human kidney organoids, we treated organoids, derived from H9 embryonic stem cells, with 5μM cisplatin for 48 hours to induce KIM-1 expression. The control and cisplatin-treated organoids were then exposed to DiI-labeled virosomes for 3 hours. Cisplatin treatment induced KIM-1 expression and KIM-1-expressing cells took up much more virosomes when compared with control organoids not exposed to cisplatin (**Fig. 4g**).

**Figure 4.**
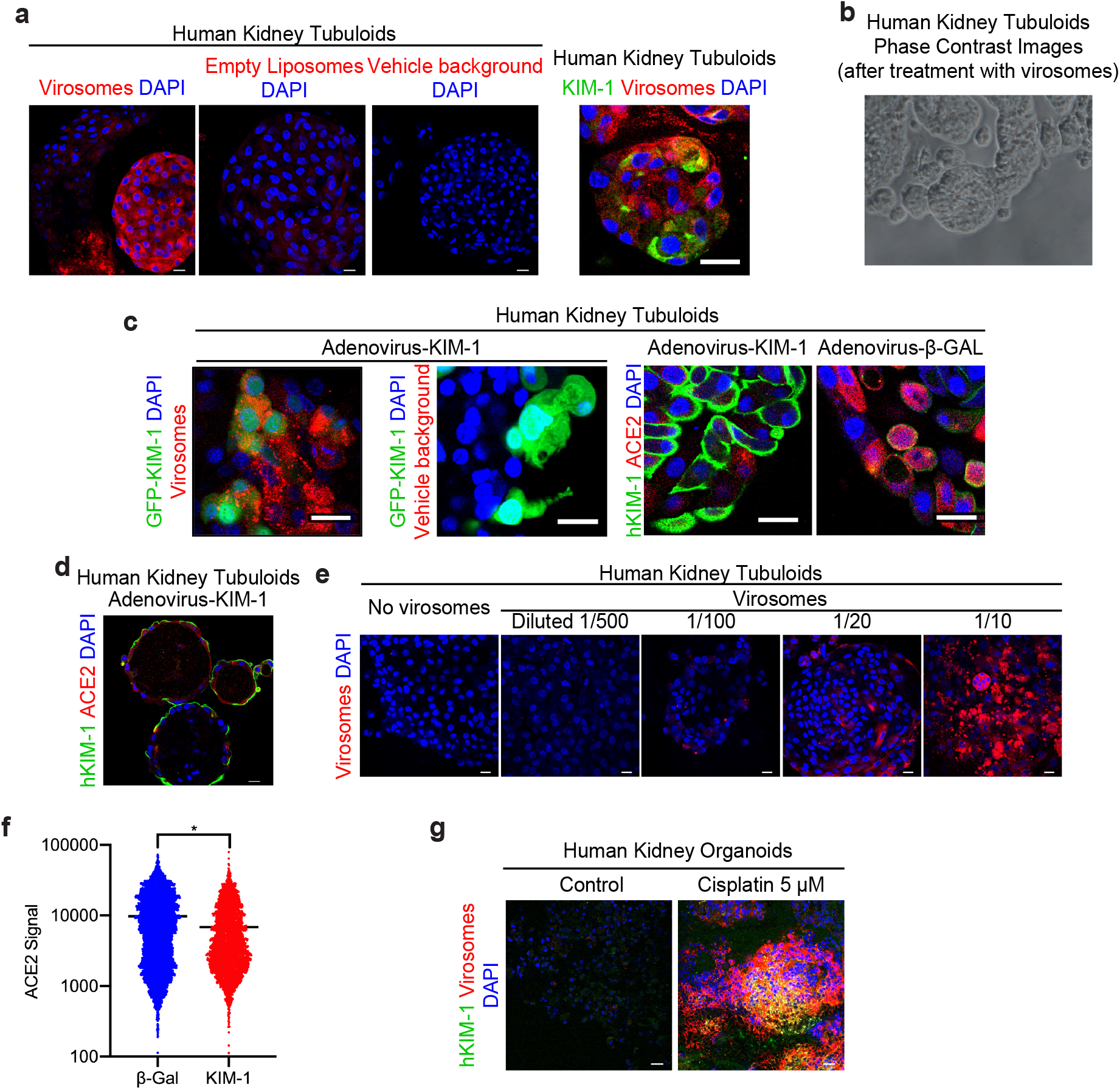
SARS-CoV-2 virosomes are internalized by human kidney tubuloids in a KIM-1-dependent manner. **(a)** Left: DiI-labeled virosome and control empty liposome internalization by human renal epithelial tubuloids. Right: Immunostaining of KIM-1 (green) in DiI-labeled virosome-treated tubuloids. Scale bars, 20 μm. **(b)** Phase contrast images of human renal epithelial tubuloids treated with SARS-CoV-2 virosomes. Scale bars, 20 μm. **(c)** Internalization assay of DiI-labeled virosomes by human renal epithelial tubuloids infected by adenovirus expressing GFP-KIM-1 (left two panels) and immunostaining of KIM-1 and ACE2 on human renal epithelial tubuloids infected by adenovirus expressing GFP-KIM-1 or control GFP-β-GAL (right two panels). Scale bars, 20 μm. **(d)** Immunostaining of KIM-1 and ACE2 in KIM-1 infected tubuloids. Scale bar, 20 μm. **(e)** Internalization assay of human renal epithelial tubuloids infected with adenovirus expressing GFP-KIM-1 and exposed to varying dilutions of DiI-labeled virosomes. Scale bars, 20 μm. **(f)** Quantification by flow cytometry of ACE2 expression of tubuloids infected with adenovirus expressing GFP-KIM-1 or control GFP-β-GAL. *p<0.0001. **(g)** Virosome uptake and KIM-1 immunostaining in human kidney organoids, untreated (Control) or treated with cisplatin (5 µM for 48 hr), and subsequently exposed to DiI-labeled virosomes. Scale bar, 20 μm.

### KIM-1 binds to SARS-CoV-2 Spike protein *in vitro*

KIM-1 binding to SARS-CoV-2 spike protein was tested by a flow cytometry-based binding assay. KIM-1-human Fc fusion protein was added to biotin-tagged SARS-CoV-2 Spike protein-bound avidin-beads to evaluate binding of KIM-1 to Spike. Bound KIM-1 was detected by flow cytometry after applying FITC-conjugated anti-human Fc antibody (**Fig. 5a and 5b**). This binding was inhibited to background levels following TW-37 addition.

**Figure 5.**
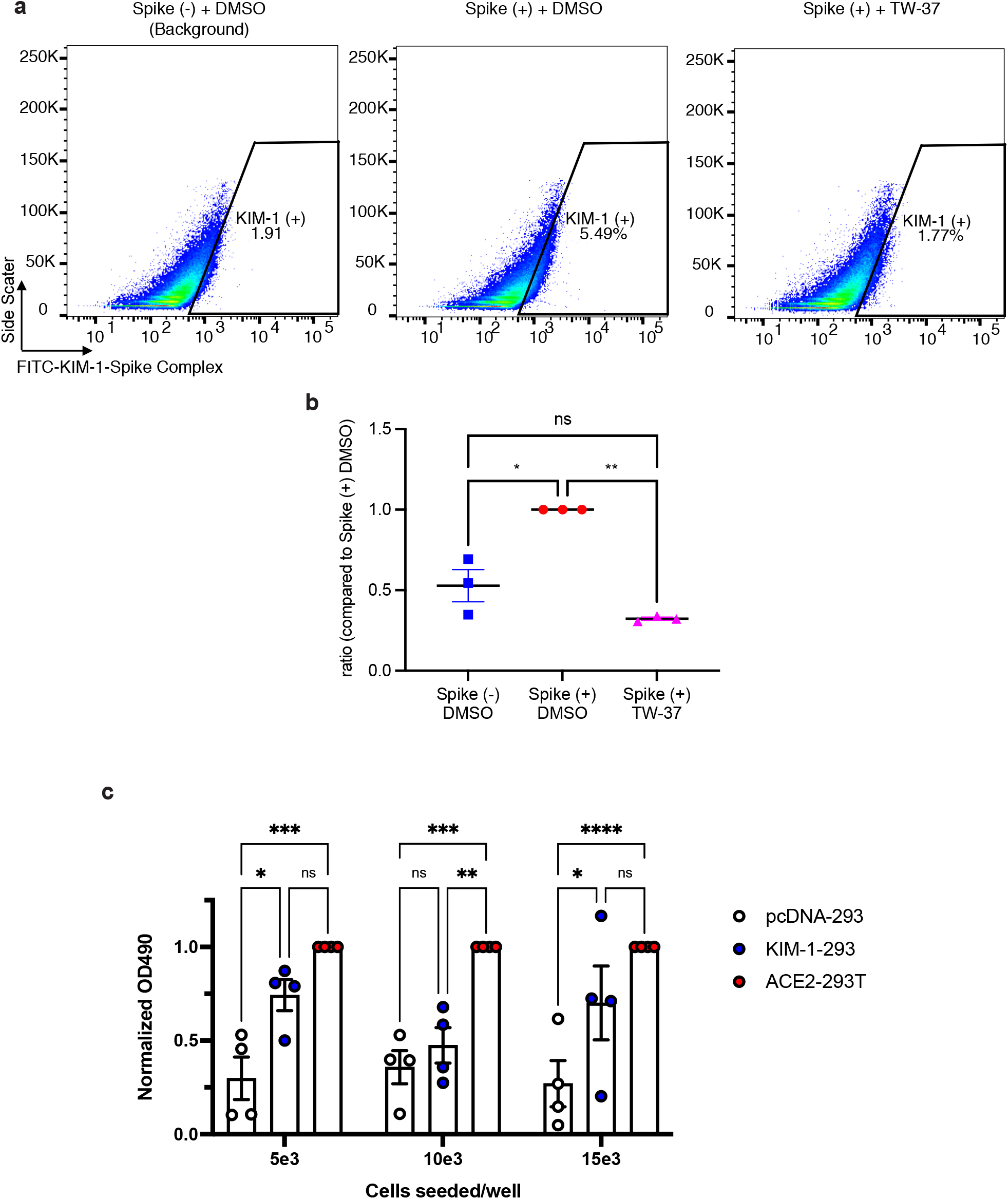
KIM-1 binds SARS-CoV-2 Spike protein *in vitro* and works as a receptor for SARS-CoV-2. **(a)** Flow cytometry-based binding assay between KIM-1 and SARS-CoV-2 Spike protein with or without TW-37. **(b)** Quantitative analysis of data in **Fig. 5a**. ^*^p=0.0029, ^**^p=0.0004. **(c)** 293 cells expressing KIM-1 without ACE2, ACE2 without KIM-1, and control pcDNA were exposed to SARS-CoV-2 and replicated virus were quantified by *in vitro* microneutralization assay. ^*^p=0.0113, ^**^p=0.0027, ^***^p=0.0001, ^****^p<0.0001.

### KIM-1 is a receptor for SARS-CoV-2

To demonstrate if KIM-1 acts as a receptor for live SARS-CoV-2, we tested whether KIM-1 enhances SARS-CoV-2 infection and replication by an *in vitro* microneutralization assay for SARS-CoV-2 ^33^. We used 293 cells expressing KIM-1 but not ACE2, 293T cells expressing ACE2 without KIM-1 and 293 cells expressing control pcDNA plasmid. KIM-1-mediated virus replication was significantly higher than the low level of replication observed with control pcDNA expressing cells and approached the level of virus replication observed with ACE2 (**Fig. 5c**). Thus, KIM-1 acts as a receptor for SARS-CoV-2 virus *in vitro*.

## DISCUSSION

We have found that KIM-1 serves as a receptor for full length replication-competent SARS-CoV-2, SARS-CoV-2 spike protein expressing pseudovirus and virosomes, and that virosome uptake and pseudovirus infection are inhibited by anti-KIM-1 antibodies or TW-37, an inhibitor for KIM-1-mediated endocytosis ^22^. Inhibition of this KIM-1-mediated uptake may be therapeutically useful in SARS-CoV-2 infection. KIM-1 protein is detectable in partially autolyzed autopsy samples due to the protein’s stability ^34^. KIM-1-expressing proximal tubules have reduced expression of ACE2, a receptor for SARS-CoV-2 ^35^, suggesting that KIM-1 may alter ACE2-mediated viral entry into cells.

Others have reported that they could not detect SARS-CoV-2 in kidneys ^36,37^. We find SARS-CoV-2 nucleocapsid protein in albeit in a small subset of proximal tubules. KIM-1 expression, however, is commonly seen in the lungs of the SARS-CoV-2-infected patients. At present, there is controversy with respect to the presence of SARS-CoV-2 nuclear capsid protein immunoreactivity in kidney tissue of subjects with documented infection with some investigators reporting its presence ^38-41^. In one study, SARS-CoV-2 RNA was reported in 38 of 63 post-mortem kidneys of patients who had SARS-CoV-2 respiratory infection ^41^. Others did not find evidence for virus in autopsy specimens ^36,42^. We found clear evidence for viral infection in one patient but the infection was quite focal and could easily have been missed in a biopsy sample. It is possible there is early infection of the kidney but then the virus is cleared by the time the kidney specimen is obtained either later in the course of illness in the patient who develops AKI or when the patient succumbs to COVID-19. This viral clearance has also been reported to occur in the lung ^43^.

Jemielity et al. reported that KIM-1 likely works as an attachment factor for SARS-CoV-1, demonstrating that KIM-1 mediates internalization of virus, but does not enhance virus replication ^19^. In contrast, KIM-1-mediated uptake of SARS-CoV-2 results in virus replication after internalization of virus. Previously, we reported that, following KIM-1-mediated phagocytosis of apoptotic cell bodies by renal epithelial cells, there was subsequent degradation involving autophagy ^16,21^. We speculate that KIM-1 can work as both an attachment factor for degradation and a receptor for SARS-CoV-2. KIM-1 may start to function as a receptor when the degradation system is highly induced due to severe infection. Specifically, SARS-CoV-2 infection-stressed renal proximal tubule epithelial cells may more strongly enhance KIM-1 expression thus promoting a stronger degradation system than may occur in lung epithelial cells. This is the speculated reason why SARS-CoV-2 infection in kidney cannot be detected as often when compared to infection in the lung.

KIM-1 is upregulated by many insults to the kidney and results in AKI. We have shown that acute upregulation of KIM-1 due to injury is protective and is likely due to an anti-inflammatory and anti-obstructive response associated with phagocytosis of apoptotic debris in the tubule ^31^. Our findings that KIM-1 is a receptor for SARS-CoV-2 indicate that KIM-1 could interact with exfoliated and virus-infected cells in the lung and kidney. Furthermore, the KIM-1 ectodomain may bind SARS-CoV-2 in the pulmonary alveoli and kidney tubular lumen and could also act as a decoy receptor. This would be reminiscent of the use of Enfuvirtide, Maraviroc and Ibalizumab for HIV where administration of agents that will bind to the virus in the circulation will compete for tissue binding ^44^.

KIM-1 facilitated endocytosis of SARS-CoV-2 virosomes in both a KIM-1- and spike protein-dependent manner. Jemielity et al. demonstrated that the phosphatidylserine binding characteristics of human KIM-1/TIM-1 mediate its activity as a SARS-CoV-1 pseudovirus receptor ^19^. It is important to recognize, however, that the virosomes we used do not contain phosphatidylserine, so the interaction of KIM-1 with this phospholipid of the virus cannot explain the KIM-1 mediated facilitation of uptake. Our binding assay confirms that the KIM-1-mediated binding interaction is with the spike protein of SARS-CoV-2.

Our recently discovered KIM-1 small molecule inhibitor, TW-37, blocked KIM-1-mediated endocytosis of SARS-CoV-2 virosomes in renal epithelial cells. TW-37 was originally discovered as a bcl-2 inhibitor in cancer drug screening, and is safe for use in animals ^45,46^. Our findings that TW-37 blocked spike protein-mediated entry of the virosome suggests a potential therapeutic role for this compound in prevention of viral internalization as an approach to anti-SARS-CoV-2 viral treatment.

In conclusion, KIM-1 is a receptor for SARS-CoV-2 via its interaction with the spike protein. KIM-1-dependent uptake of SARS-CoV-2 by lung and kidney cells can be inhibited by anti-KIM-1 antibodies and TW-37. This may have important implications for viral entry, initiation of virus replication, and/or inactivation of the virus through viral degradation or decoy function in the kidney and respiratory mucosa. Targeted treatment directed at the KIM-1-SARS-CoV-2 interaction may be both therapeutic and prophylactic for this devastating disease that occurs secondary to infection.

## Data Availability

All the relevant data are available from the corresponding authors upon reasonable request.

## Acknowledgements

This work was supported by grants from the National Institute of Health/NCATS/NIDDK UH3TR002155 (J.V.B. & L.P.L.); NIDDK 2R01DK072381 (J.V.B. & L.P.L.), R37DK039773 (J.V.B.) and Postdoctoral Fellowship from Uehara Memorial Foundation (to Y.M.), the Overseas Research Fellowships from Japan Society for the Promotion of Science (to Y.M.) and the Canadian Institutes of Health Research (to G.A.D. and J.D.D.). The authors would like to thank Zi-Fu Wang for help with binding experiments and thank Bing Chen and Youngfei Cai for providing purified spike ectodomain. We thank Drs. Huihui Mou and Michael Farzan for providing ACE2-293T cells. We thank Dr. Bing Chen for providing a stable Expi293F cells expressing S.dTM.PP.

## Author contributions

Y.M., C.F., T.I., K.S., M.M., P.A., N.N.L., K.M.P.D., R.F.P., A.W., S.G.H., M.S., and M.L.N. performed experiments, collected and analyzed data, and wrote the manuscript. R.F.P. and A.W. obtained human lung and kidney samples and helped to interpret the pathology. L.P.L supported S.G.H.’s and M.S.’s experiment. G.A.D. and J.D.D supported C.F.’s SARS-CoV-2 experiments. Y.M. and J.V.B. developed experimental strategy, supervised the project, and edited the manuscript. All authors discussed the results and implications and commented on the manuscript.

## Additional Information

### Correspondence and requests for materials

should be addressed to Y.M. and J.V.B.

## Methods

### Human post-mortem samples and biopsy samples

Human postmortem lung and kidney samples from COVID-19 patients and human kidney biopsy samples from COVID-19-associated AKI patients were obtained from clinically indicated pathological autopsy and kidney biopsies in Brigham and Women’s Hospital in Boston, US. The protocol was approved by the Institutional Review Board of the Ethics Committee of Partners Healthcare.

### Immunofluorescence staining of paraffin sections

Human paraffin sections of postmortem samples or biopsy samples were deparaffinized with xylene, ethanol, 2% hydrogen peroxide in methanol to ablate peroxidase activity in a microwave oven. The sections were blocked with 3% BSA-PBS and were incubated with primary antibodies for 1 hour at room temperature. After washing with PBS, sections were incubated with secondary antibodies for 30 minutes. Vectashield (Vector Laboratories, Burlingame, CA) containing DAPI (12.5 μg/mL) was applied and the slides were cover-slipped. All images were obtained by confocal microscopy (C1 Eclipse from Nikon).

### Immunohistochemistry staining of SARS nucleocapsid protein in paraffin sections

Human paraffin sections of postmortem samples were deparaffinized with xylene, ethanol, 2% hydrogen peroxide in methanol to ablate peroxidase activity in a microwave oven. The sections were blocked with Avidin/Biotin Blocking Kit (Vector Laboratories, Burlingame, CA) and were incubated with primary antibody (anti-SARS nucleocapsid protein) for 1 hour at room temperature. Nucleocapsid protein was detected by using Vectastain Elite ABC kit (for mouse IgG) with DAB Substrate (Vector Laboratories, Burlingame, CA) kit for peroxidase staining. The sections were counterstained with hematoxylin and then mounted with cover slips using Permount (Sigma-Aldrich, St. Louis, MO).

### Mouse ischemia reperfusion injury (IRI)

C57BL/6 mice were purchased from Charles River Laboratories. All mouse work was performed in accordance with the animal use protocol approved by the Institutional Animal Care and User Committee of the Harvard Medical School. Mice aged 8-12 weeks and weighing 20-22 g were subjected to IRI according to procedures as described previously ^16,47^. Briefly, both kidneys were exposed by flank incisions, and the renal pedicles were clamped for 25 min at 37°C. After surgery, 1 mL of warm saline (37°C) was injected intraperitoneally for volume supplementation. Sham operations were performed by exposing both kidneys without clamping of renal pedicles. Kidney tissue was collected at 24 hours or 48 hours post IRI.

### Cell culture experiments

Human primary renal epithelial cells were obtained from the uninvolved parts of kidneys removed for nephrectomy on the renal cell carcinoma or other kidney cancers in Brigham and Women’s Hospital in Boston, US, by modifying a previously established protocol ^21^. The protocol was approved by the Institutional Review Board of the Ethics Committee of Partners Healthcare. Mouse primary lung epithelial cells were obtained and cultured by using the same protocol ^21^. Briefly, human renal cortex was minced, or mouse lung was taken out after sacrificed and was minced. They were digested with collagenase (0.5 mg/mL) in DMEM/F12 50:50 media for 40 minutes. The enzyme reaction was deactivated with fetal bovine serum (FBS). After gravity sedimentation for 2 minutes, supernatant was discarded. The remaining sample was washed 2 times in media and tissues were resuspended in primary cell culture medium (DMEM/F12 with BSA, transferrin, insulin, selenium, hydrocortisone, and epidermal growth factor (EGF)). The epithelial cells were cultured for 7 to 14 days before being used for experiments.

Mutant mouse KIM-1 expression plasmid (WFAA or AAND in phosphatidylserine-binding motif, WFND (amino acid residues 112-115 in Ig domain)) were generated by site-directed mutagenesis as previously described ^16^. LLC-PK1 cells stably expressing human or mouse KIM-1 were generated by transfecting LLC-PK1 cells with human or mouse KIM-1 full-length cDNA in the pcDNA vector ^16,21^. LLC-PK1 cells stably expressing empty pcDNA were used as controls.

### Virosomes assembly and purification

SARS-CoV-2 spike ectodomain was generated by using plasmid S.dTM.PP which was previously reported ^48^, or purchased only for **Fig. 1b** (Biotinylated SARS-CoV-2 (COVID-19) S1 protein, His,Avitag™ (MALS verified), AcroBiosystems, Newark, DE). We obtained a stable Expi293F cells expressing S.dTM.PP from Dr. Bing Chen at Boston Children’s Hospital. Purified SARS-CoV-2 spike ectodomain with deletion of the furin cleavage site, two proline mutations, a foldon trimerization domain and a C terminal His tag was used in the virosome assembly. Chloroform lipid stocks were mixed to accomplish the following ratio 15% 1,2-dioleoyl-sn-glycero-3-[(N-(5-amino-1-carboxypentyl) iminodiacetic acid) succinyl] (nickel, Ni, salt) (18:1DGS nickel-nitrilotriacetic acid (NTA) Ni)/51% 2-oleoyl-1-pamlitoyl-sn-glycero-3-phosphocholine (POPC)/34% 2-oleoyl-1-pamlitoyl-sn-glycero-3-glycerol (POPG). The mixture was dried under argon stream and stored in a desiccator overnight. The next day the mixture was resuspended in PBS buffer to achieve a final lipid concentration of 10 mM, the resulting milky solution was repeatedly pushed through a 0.1 μm filter membrane using an extruder until the solution was turned clear. The resulting liposome solution was calculated to have a concentration of approximately 100 nM. Liposomes were mixed with the His-tagged Spike ectodomain to achieve a ratio of 40:1 (spike-trimer: liposome). The mixture was incubated for 30 min on 4 °C to allow the spike His-tag to bind to the Ni-NTA lipids. Free spike protein was separated from spike loaded liposomes using a Superose 6 10/300 size exclusion chromatography column.

### Internalization assay of virosomes and control empty liposomes

Cells were seeded onto 8-well chamber slide at a density of 1-1.5 × 10^5^ cells/well (LLC-PK1 cells stably expressing human KIM-1 or empty pcDNA, A549 cells or mouse primary lung epithelial cells). After the cells reached confluency, the cells were treated with 10 μM TW-37 or control DMSO for 30 minutes, or with 10 μM TW-37 or control DMSO in DMEM/F-12 media, and 0.025 mg/mL anti-KIM-1 mouse IgG (3F4) and 0.025 mg/mL anti KIM-1 mouse IgG (AKG7), or 0.05 mg/mL control mouse IgG in DMEM/F-12 media for 1 hour as pretreatment. Then virosomes or control empty liposomes were added at a ratio of 1:10 to media, and cells were incubated for 1.5 hours at 37 ºC. After exposure to liposomes, the cells were washed with PBS and fixed with 4% paraformaldehyde (PFA)-PBS. Immunofluorescence staining of the cells was performed according to each experiment. For quantification of internalization from images taken by confocal microscopy, virosome-positive areas were measured by ImageJ and analyzed statistically.

For quantification of internalization by flow cytometry, A549 cells were seeded onto 12-well plates at a density of 4 × 10^5^ cells/well. After the cells become mostly confluent, the cells were treated with 10 μM TW-37 or control DMSO in DMEM/F-12 media, and 0.025 mg/mL anti-KIM-1 mouse IgG (3F4) and 0.025 mg/mL anti KIM-1 mouse IgG (AKG7), or 0.05 mg/mL control mouse IgG in DMEM/F-12 media for 1 hour as pretreatment. Then virosomes or control empty liposomes were added at a ratio of 1:10 to media, and cells were incubated for 1.5 hours at 37 ºC. After exposure to liposomes, the cells were washed with PBS and detached with 0.25% Trypsin/0.1% EDTA, fixed with 4% paraformaldehyde/5% FBS in PBS, and then immunostained with anti-KIM-1 and anti-ACE2 antibodies. Cells were analyzed by FACS Canto II (BD Biosciences). Data were analyzed using FLOWJO (FLOWJO).

### Immunofluorescence staining of cells

After fixation with 4% PFA-PBS, the cells were permeabilized with 0.1% Triton X-100-PBS and blocked with 3% BSA-PBS for 30 minutes. Primary antibodies were applied and the slides were incubated for 1 hour at room temperature or overnight at 4 ºC. After washing with PBS, the slides were exposed to secondary antibodies and incubated for 30 minutes at room temperature. All images were obtained by confocal microscopy (C1 Eclipse from Nikon).

### Antibodies

In immunofluorescence staining, primary antibodies against the following proteins were used: human KIM-1 (goat, 1:200, AF1750; R&D systems Inc, Minneapolis, MN); human KIM-1 (mouse, 1:1, AKG7 ^20^, developed in collaboration with BIOGEN Inc. Cambridge, MA); mouse KIM-1 (goat, 1:200, AF1817; R&D systems Inc, Minneapolis, MN); SARS Coronavirus Nucleocapsid (rabbit, 1:200, PA1-41098; Invitrogen, Thermo Fisher Scientific, Waltham, MA); ACE2 (rabbit, 1:200, ab15348; Abcam, Cambridge, MA); Prosurfactant Protein C (rabbit, 1:200, ab90716; Abcam, Cambridge, MA); pan-cytokeratin (mouse, 1:200, Sigma-Aldrich, St. Louis, MO). Secondary antibodies were either FITC-, Cy3- or Cy5-conjugated (Jackson ImmunoResearch Inc., West Grove, PA). For western blotting, primary antibodies against the following proteins were used: ACE2 (1:1000, as used in immunofluorescence staining); human KIM-1 cytoplasmic domain (rabbit, 1:1000, #195, developed in collaboration with BIOGEN Inc. Cambridge, MA ^49^); ERK1/2 (goat, 1:1000, Santa Cruz Biotechnology, Dallas, TX). As secondary antibodies, HRP-conjugated anti-rabbit IgG and anti-goat IgG (Dako, Denmark) were used. For inhibition of KIM-1 by antibodies, purified anti-human KIM-1 IgG (both mouse monoclonal AKG7 and 3F4, developed by our group) or anti-mouse KIM-1 IgG (AF1817) were used. Control mouse IgG was purchased from Jackson ImmunoResearch Inc. (West Grove, PA).

### Western blotting

Cells and kidneys were lysed and protein was purified as previously described ^11^. Bands were visualized by chemiluminescence (Western Lightning, PerkinElmer, Waltham, MA).

### Human alveoloids

A549 cells were cultured in ultra-low attachment plates with 10% FBS-DMEM. After around 8 weeks, alveoloids were used for experiments.

### Human renal tubuloids

The manuscript on the protocol to make human renal tubuloids is in preparation. Briefly, human primary renal epithelial cells were cultured on ultra-low attachment plates with 5% FBS-RPMI. After 2-3 days incubation, Matrigel was added and media was changed to 5% FBS-Advanced RPMI containing EGF, bFGF and HGF. Media was changed once or twice a week. The tubuloids are ready for use after 2 weeks. Adenoviruses for expression of human KIM-1 or control β-galactosidase (β-GAL) were produced and stored as described previously ^49^.

### Human kidney organoids. Human kidney organoids were derived from H9 embryonic pluripotent stem cells (hESC)

Organoids were generated following the methods in our previous study with modifications ^50^. Briefly, H9 hESC were grown in mTeSR1 (Stemcell Technologies). Cells were dissociated and plated for differentiation process. Plated cells were differentiated utilizing the drugs and factors outlined in the previous study including Rock Inhibitor (Tocris), CHIR (Tocris), and FGF9 (R&D Systems) in Advanced RPMI media (Thermo Fischer) and grown to maturity in 96 well ultra-low attachment plate (Corning). The human kidney organoids were harvested for use after day 30 from the beginning of differentiation.

### *In vitro* binding assay

10 μL of Biotinylated 2019-nCoV S1 protein, His, Avitag, 25 μL of FluoSpheres NeutrAvidin-Labeled Microspheres, 1.0 µm, nonfluorescent, 1% solids (Thermo Fisher Scientific, MA), and 250 μL PBS were mixed and incubated for 30 minutes at room temperature on a belly dancer. The microspheres were washed by PBS, then incubated with or without 10 μM TW-37 at room temperature for 15 minutes. 10 μL of Recombinant Human TIM-1/KIM-1/HAVCR1 Fc Chimera Protein (0.5 mg/mL, 9319-TM; R&D systems Inc, Minneapolis, MN) was added into the mixture. After incubation at room temperature for 1 hour on a belly dancer, the microspheres were washed by PBS. FITC-conjugated goat-anti-human Fc antibody was applied and the mixture was incubated at room temperature for 30 minutes. After washing with PBS, the microspheres were analyzed by FACS Canto II (BD Biosciences). Data were analyzed using FLOWJO (FLOWJO).

### *In vitro* microneutralization assay for SARS-CoV-2

An *in vitro* microneutralization assay for SARS-CoV-2 was conducted using a previously defined protocol ^33^. Briefly, 293 cells stably expressing KIM-1 or control pcDNA, or 293T cells stably expressing ACE2 were seeded on a 96-well plate and infected with replication-competent SARS-CoV-2 USA-WA1/2020 virus strain in Biosafety Level 3 (BSL-3) laboratory. After 24 hours, SARS-CoV-2 infection was quantified using mouse anti-SARS-CoV-2 nucleocapsid protein and anti-mouse IgG HRP antibodies and an OPD developing solution. The optical density at 490 nm was measured using a microplate reader and served as the assay readout.

### Pseudovirus binding and entry into LLC-PK1 co-cultured cells

KIM-1-PK1 cells and control pcDNA-PK1 cells were cultured separately in 12-well plates before the co-culture. KIM-1-PK1 was stained with an orange-color fluorescent dye (CellTracker Orange CMRA, Thermofisher Scientific, MA) for 30min before detached. KIM-1-PK1 cells and pcDNA-PKA cells were single-cell dissociated with Trypsin/EDTA and mixed as 2:1 ratio in plating. At 90% confluency, the cells were transduced with coronavirus-based SARS-CoV-2 pseudovirus (CoV2-01, SARS-CoV-2-S(GFP), Virongy, VA) for 12 hours, refreshed with media, and imaged 1.5 days after transfection.

### Cell lines and reagents

LLC-PK1 cell lines, A549 cell lines and 293 cell lines were obtained from the ATCC. FBS, DMEM, and DMEM/F-12 were from Cellgro (Manassas, VA). Cell lines stably expressing pcDNA or KIM-1 were generated as described previously ^21^. ACE2-293T cells were kindly provided by Drs. Huihui Mou and Michael Farzan ^51^. TW-37 was purchased from Selleck Chemicals (Houston, TX). For labeling the virosomes and empty liposomes, CellTracker CM-DiI was purchased from Molecular Probes, Inc (Eugene, OR).

### Statistical methods

Data are reported as mean ± standard error of the mean (SEM). Number of samples assayed in each experiment is indicated in the Figure Legends. Tukey-Kramer Multiple Comparisons Test was used for multiple comparisons. p<0.05 was considered to represent a statistically significant difference. Prism 8 (GraphPad Software, LLC) was used for all the statistical analysis.

## Resource Availability

### Lead Contact

Further information and requests for resources and reagents should be directed to and will be fulfilled by the Lead Contact, Joseph V. Bonventre.

### Material Availability

This study did not generate new unique reagents.

### Data and Code Availability

All other data are available from the Lead Contact on reasonable request.

## References

1 Naming the coronavirus disease (COVID-19) and the virus that causes it, <https://www.who.int/emergencies/diseases/novel-coronavirus-2019/technical-guidance/naming-the-coronavirus-disease-(covid-2019)-and-the-virus-that-causes-it> (2020).

2 Hui, D. S. et al. The continuing 2019-nCoV epidemic threat of novel coronaviruses to global health - The latest 2019 novel coronavirus outbreak in Wuhan, China. Int J Infect Dis 91, 264–266, doi:10.1016/j.ijid.2020.01.009 (2020).

3 WHO Director-General’s opening remarks at the media briefing on COVID-19 - 11 March 2020, <https://www.who.int/dg/speeches/detail/who-director-general-s-opening-remarks-at-the-media-briefing-on-covid-1911-march-2020> (2020).

4 Coronavirus COVID-19 Global Cases by the Center for Systems Science and Engineering (CSSE) at Johns Hopkins University (JHU), <https://coronavirus.jhu.edu/map.html> (2020).

5 Li, Z. et al. Caution on Kidney Dysfunctions of 2019-nCoV Patients. medRxiv, 2020.2002.2008.20021212, doi:10.1101/2020.02.08.20021212 (2020).

6 Chen, N. et al. Epidemiological and clinical characteristics of 99 cases of 2019 novel coronavirus pneumonia in Wuhan, China: a descriptive study. Lancet 395, 507–513, doi:10.1016/S0140-6736(20)30211-7 (2020).

7 Guan, W. J. et al. Clinical Characteristics of Coronavirus Disease 2019 in China. N Engl J Med 382, 1708–1720, doi:10.1056/NEJMoa2002032 (2020).

8 Cheng, Y. et al. Kidney impairment is associated with in-hospital death of COVID-19 patients. medRxiv, 2020.2002.2018.20023242, doi:10.1101/2020.02.18.20023242 (2020).

9 Chen, T. et al. Clinical characteristics of 113 deceased patients with coronavirus disease 2019: retrospective study. BMJ 368, m1091, doi:10.1136/bmj.m1091 (2020).

10 Kaplan, G. et al. Identification of a surface glycoprotein on African green monkey kidney cells as a receptor for hepatitis A virus. Embo j 15, 4282–4296 (1996).

11 Ichimura, T. et al. Kidney injury molecule-1 (KIM-1), a putative epithelial cell adhesion molecule containing a novel immunoglobulin domain, is up-regulated in renal cells after injury. The Journal of biological chemistry 273, 4135–4142 (1998).

12 Ichimura, T., Hung, C. C., Yang, S. A., Stevens, J. L. & Bonventre, J. V. Kidney injury molecule-1: a tissue and urinary biomarker for nephrotoxicant-induced renal injury. American journal of physiology. Renal physiology 286, F552–563, doi:10.1152/ajprenal.00285.2002 (2004).

13 Vaidya, V. S. et al. Kidney injury molecule-1 outperforms traditional biomarkers of kidney injury in preclinical biomarker qualification studies. Nat Biotechnol 28, 478–485, doi:10.1038/nbt.1623 (2010).

14 Takasu, O. et al. Mechanisms of cardiac and renal dysfunction in patients dying of sepsis. Am J Respir Crit Care Med 187, 509–517, doi:10.1164/rccm.201211-1983OC (2013).

15 Sabbisetti, V. S. et al. Blood kidney injury molecule-1 is a biomarker of acute and chronic kidney injury and predicts progression to ESRD in type I diabetes. J Am Soc Nephrol 25, 2177–2186, doi:10.1681/ASN.2013070758 (2014).

16 Brooks, C. R. et al. KIM-1-/TIM-1-mediated phagocytosis links ATG5-/ULK1-dependent clearance of apoptotic cells to antigen presentation. Embo j 34, 2441–2464, doi:10.15252/embj.201489838 (2015).

17 Kondratowicz, A. S. et al. T-cell immunoglobulin and mucin domain 1 (TIM-1) is a receptor for Zaire Ebolavirus and Lake Victoria Marburgvirus. Proc Natl Acad Sci U S A 108, 8426–8431, doi:10.1073/pnas.1019030108 (2011).

18 Meertens, L. et al. The TIM and TAM families of phosphatidylserine receptors mediate dengue virus entry. Cell Host Microbe 12, 544–557, doi:10.1016/j.chom.2012.08.009 (2012).

19 Jemielity, S. et al. TIM-family proteins promote infection of multiple enveloped viruses through virion-associated phosphatidylserine. PLoS Pathog 9, e1003232, doi:10.1371/journal.ppat.1003232 (2013).

20 Bailly, V. et al. Shedding of kidney injury molecule-1, a putative adhesion protein involved in renal regeneration. The Journal of biological chemistry 277, 39739–39748, doi:10.1074/jbc.M200562200 (2002).

21 Ichimura, T. et al. Kidney injury molecule-1 is a phosphatidylserine receptor that confers a phagocytic phenotype on epithelial cells. The Journal of clinical investigation 118, 1657–1668, doi:10.1172/jci34487 (2008).

22 Mori, Y. et al. KIM-1 mediates fatty acid uptake by renal tubular cells to promote progressive diabetic kidney disease. Cell metabolism 33, 1042–1061 e1047, doi:10.1016/j.cmet.2021.04.004 (2021).

23 Du, L. et al. The spike protein of SARS-CoV--a target for vaccine and therapeutic development. Nat Rev Microbiol 7, 226–236, doi:10.1038/nrmicro2090 (2009).

24 Krueger, D. K., Kelly, S. M., Lewicki, D. N., Ruffolo, R. & Gallagher, T. M. Variations in disparate regions of the murine coronavirus spike protein impact the initiation of membrane fusion. J Virol 75, 2792–2802, doi:10.1128/jvi.75.6.2792-2802.2001 (2001).

25 Lewicki, D. N. & Gallagher, T. M. Quaternary structure of coronavirus spikes in complex with carcinoembryonic antigen-related cell adhesion molecule cellular receptors. The Journal of biological chemistry 277, 19727–19734, doi:10.1074/jbc.M201837200 (2002).

26 Huang, Y. H. et al. CEACAM1 regulates TIM-3-mediated tolerance and exhaustion. Nature 517, 386–390, doi:10.1038/nature13848 (2015).

27 Thomas, L. J. et al. Development of a Novel Antibody-Drug Conjugate for the Potential Treatment of Ovarian, Lung, and Renal Cell Carcinoma Expressing TIM-1. Mol Cancer Ther 15, 2946–2954, doi:10.1158/1535-7163.MCT-16-0393 (2016).

28 Zheng, X., Xu, K., Chen, L., Zhou, Y. & Jiang, J. Prognostic value of TIM-1 expression in human non-small-cell lung cancer. J Transl Med 17, 178, doi:10.1186/s12967-019-1931-2 (2019).

29 Harcourt, J. et al. Severe Acute Respiratory Syndrome Coronavirus 2 from Patient with 2019 Novel Coronavirus Disease, United States. Emerg Infect Dis 26, doi:10.3201/eid2606.200516 (2020).

30 Xiao, S. et al. Defect in regulatory B-cell function and development of systemic autoimmunity in T-cell Ig mucin 1 (Tim-1) mucin domain-mutant mice. Proc Natl Acad Sci U S A 109, 12105–12110, doi:10.1073/pnas.1120914109 (2012).

31 Yang, L. et al. KIM-1-mediated phagocytosis reduces acute injury to the kidney. The Journal of clinical investigation 125, 1620–1636, doi:10.1172/jci75417 (2015).

32 Abcam. Anti-ACE2 antibody (ab15348), <https://www.abcam.com/ace2-antibody-ab15348.html> (2021).

33 Amanat, F. et al. An In Vitro Microneutralization Assay for SARS-CoV-2 Serology and Drug Screening. Curr Protoc Microbiol 58, e108, doi:10.1002/cpmc.108 (2020).

34 Yin, W., Zhang, P. L., Macknis, J. K., Lin, F. & Bonventre, J. V. Kidney injury molecule-1 identifies antemortem injury in postmortem adult and fetal kidney. American journal of physiology. Renal physiology 315, F1637–F1643, doi:10.1152/ajprenal.00060.2018 (2018).

35 Hoffmann, M. et al. SARS-CoV-2 Cell Entry Depends on ACE2 and TMPRSS2 and Is Blocked by a Clinically Proven Protease Inhibitor. Cell 181, 271–280 e278, doi:10.1016/j.cell.2020.02.052 (2020).

36 Sharma, P. et al. COVID-19-Associated Kidney Injury: A Case Series of Kidney Biopsy Findings. J Am Soc Nephrol, doi:10.1681/ASN.2020050699 (2020).

37 Kudose, S. et al. Kidney Biopsy Findings in Patients with COVID-19. J Am Soc Nephrol 31, 1959–1968, doi:10.1681/ASN.2020060802 (2020).

38 Farkash, E. A., Wilson, A. M. & Jentzen, J. M. Ultrastructural Evidence for Direct Renal Infection with SARS-CoV-2. J Am Soc Nephrol 31, 1683–1687, doi:10.1681/ASN.2020040432 (2020).

39 Su, H. et al. Renal histopathological analysis of 26 postmortem findings of patients with COVID-19 in China. Kidney international 98, 219–227, doi:10.1016/j.kint.2020.04.003 (2020).

40 Puelles, V. G. et al. Multiorgan and Renal Tropism of SARS-CoV-2. N Engl J Med 383, 590–592, doi:10.1056/NEJMc2011400 (2020).

41 Braun, F. et al. SARS-CoV-2 renal tropism associates with acute kidney injury. The Lancet, doi:10.1016/S0140-6736(20)31759-1 (2020).

42 Santoriello, D. et al. Postmortem Kidney Pathology Findings in Patients with COVID-19. J Am Soc Nephrol, doi:10.1681/ASN.2020050744 (2020).

43 Schaefer, I. M. et al. In situ detection of SARS-CoV-2 in lungs and airways of patients with COVID-19. Mod Pathol, doi:10.1038/s41379-020-0595-z (2020).

44 Henrich, T. J. & Kuritzkes, D. R. HIV-1 entry inhibitors: recent development and clinical use. Curr Opin Virol 3, 51–57, doi:10.1016/j.coviro.2012.12.002 (2013).

45 Ahn, C. H. et al. Antitumor effect of TW-37, a BH3 mimetic in human oral cancer. Lab Anim Res 35, 27, doi:10.1186/s42826-019-0028-7 (2019).

46 Lei, S. et al. The preclinical analysis of TW-37 as a potential anti-colorectal cancer cell agent. PLoS One 12, e0184501, doi:10.1371/journal.pone.0184501 (2017).

47 Park, K. M. et al. Inducible nitric-oxide synthase is an important contributor to prolonged protective effects of ischemic preconditioning in the mouse kidney. The Journal of biological chemistry 278, 27256–27266, doi:10.1074/jbc.M301778200 (2003).

48 Yu, J. et al. DNA vaccine protection against SARS-CoV-2 in rhesus macaques. Science 369, 806–811, doi:10.1126/science.abc6284 (2020).

49 Zhang, Z., Humphreys, B. D. & Bonventre, J. V. Shedding of the urinary biomarker kidney injury molecule-1 (KIM-1) is regulated by MAP kinases and juxtamembrane region. J Am Soc Nephrol 18, 2704–2714 (2007).

50 Morizane, R. et al. Nephron organoids derived from human pluripotent stem cells model kidney development and injury. Nat Biotechnol 33, 1193–1200, doi:10.1038/nbt.3392 (2015).

51 Mou, H. et al. Mutations from bat ACE2 orthologs markedly enhance ACE2-Fc neutralization of SARS-CoV-2. bioRxiv, doi:10.1101/2020.06.29.178459 (2020).

