## Supplemental Figures for "KIM-1/TIM-1 is a Receptor for SARS-CoV-2 in Lung and Kidney"

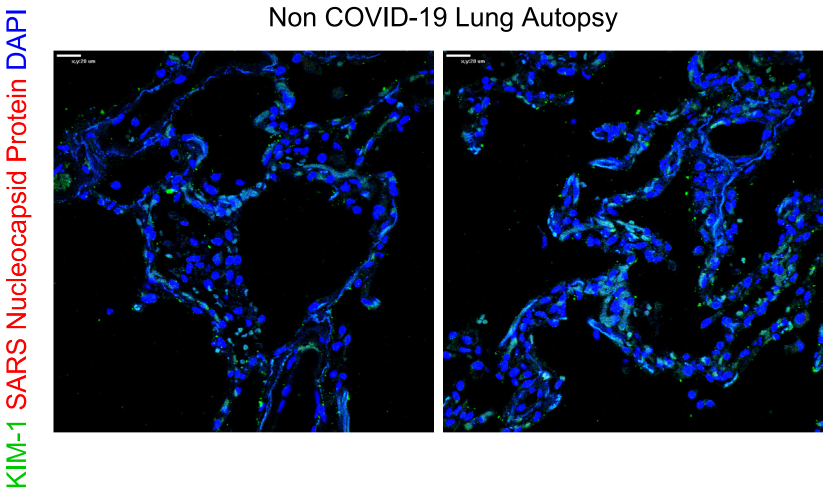


**Supplementary Figure 1. Non COVID-19 lungs expresses very low amount of KIM-1.** Immunostaining of KIM-1 and SARS Nucleocapsid Protein. Scale bars: 20 μm.

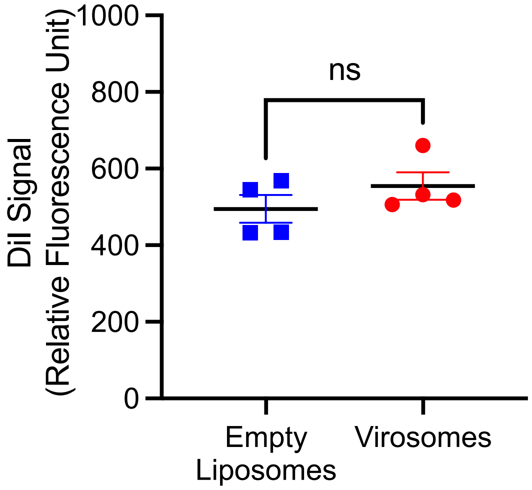


**Supplementary Figure 2. No significant difference was observed between DiI signal from labeled empty liposomes and labeled virosomes.** DiI signal was detected by using fluorescence plate reader. p=0.1427.

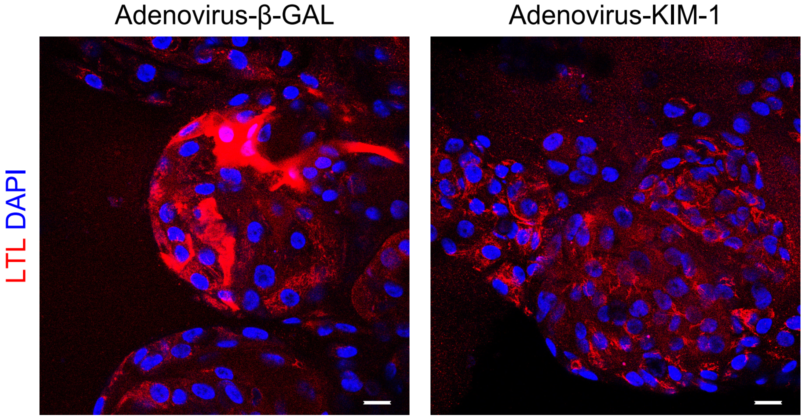


**Supplementary Figure 3. Adenovirus-KIM-1 infection decreases Lotus tetragonolobus lectin (LTL) expression when compared with Adenovirus-β-gal infection of tubuloids.**

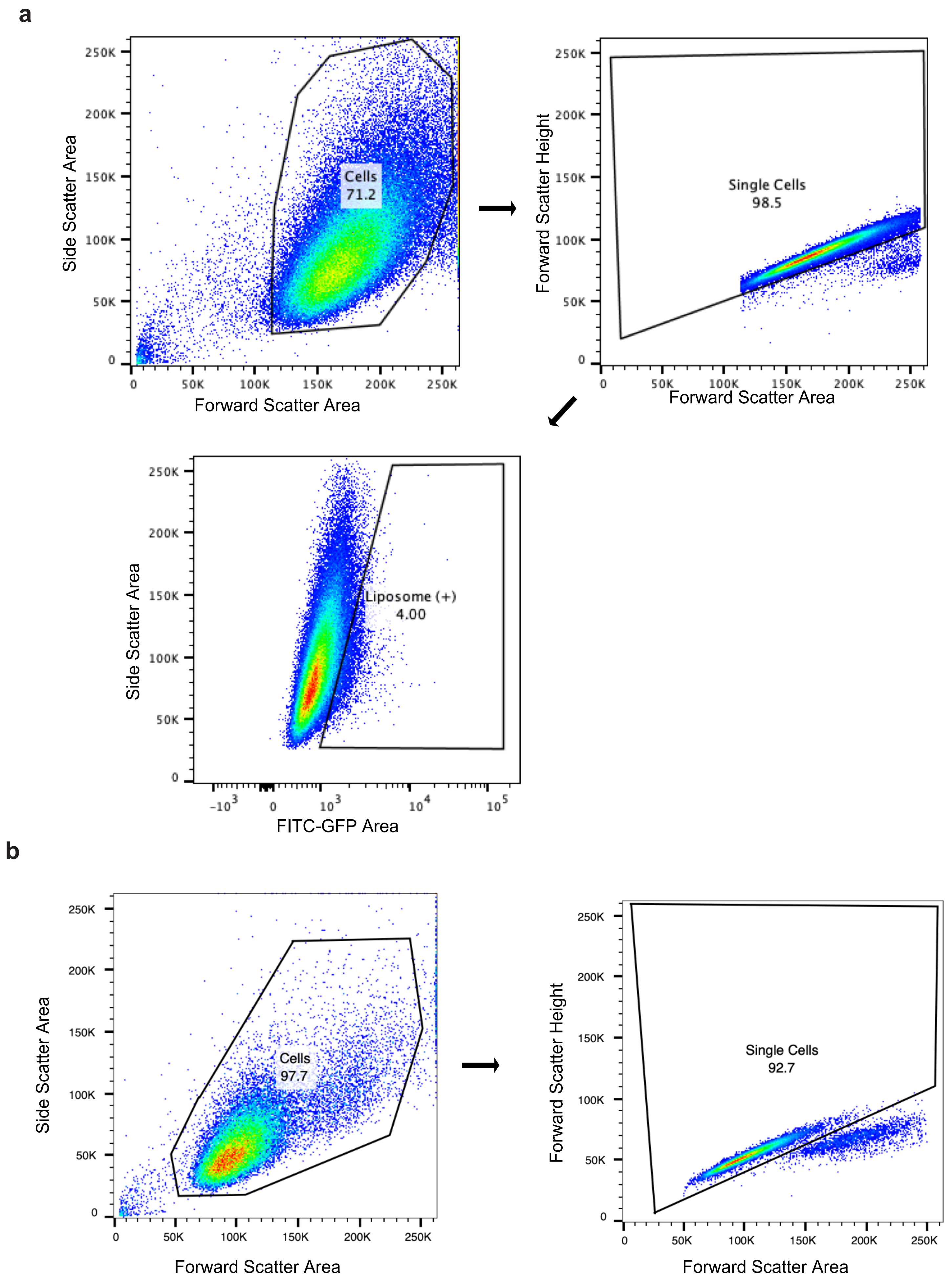


**Supplementary Figure 4. Gating Strategy for flow cytometry for cells.** (**a**) Gating strategy on the analysis of virosome (or control liposome)-treated A549 cells for **Fig. 2b**. (**b**) Gating strategy on the analysis of virosome-treated A549 cells for **Fig. 2e**.

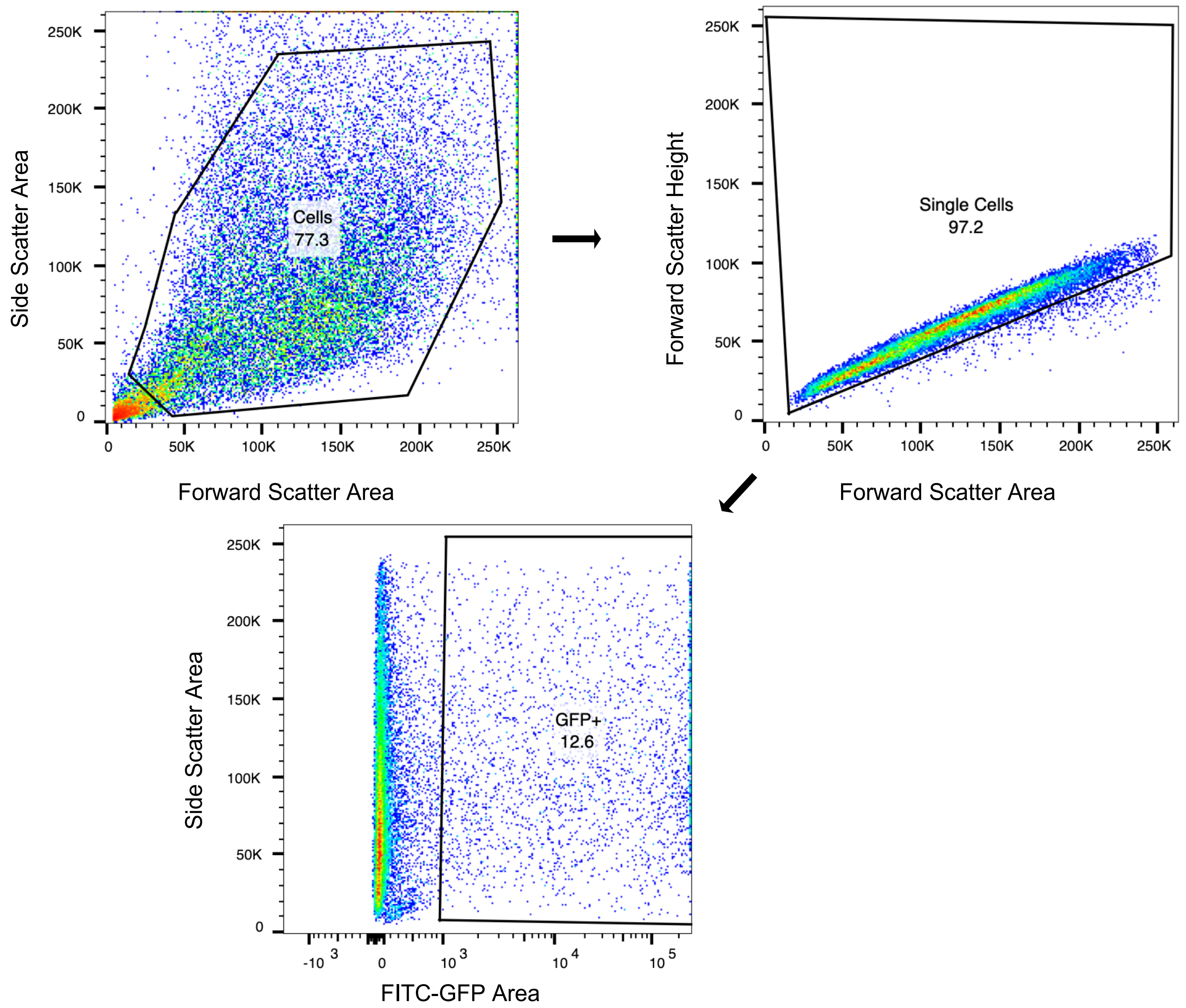


**Supplementary Figure 5. Gating Strategy for flow cytometry for tubuloids.** Gating strategy on the analysis of virosome-treated and digested human renal tubuloids (**Fig.4f**).

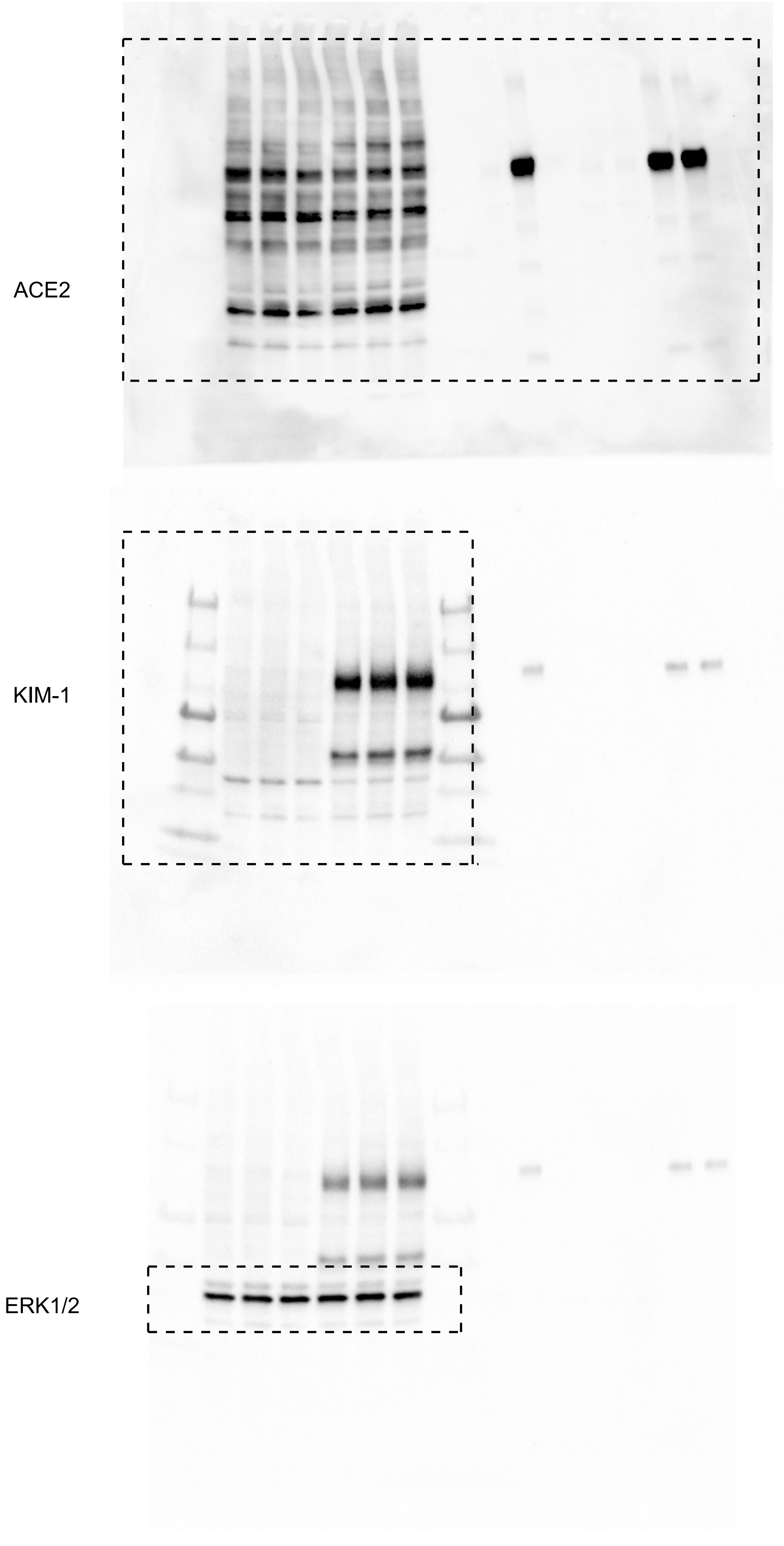


**Supplementary Figure 6. Whole membrane images of western blotting in Fig. 3c.**
The areas surrounded by the dotted line are shown as **Fig. 3c**.

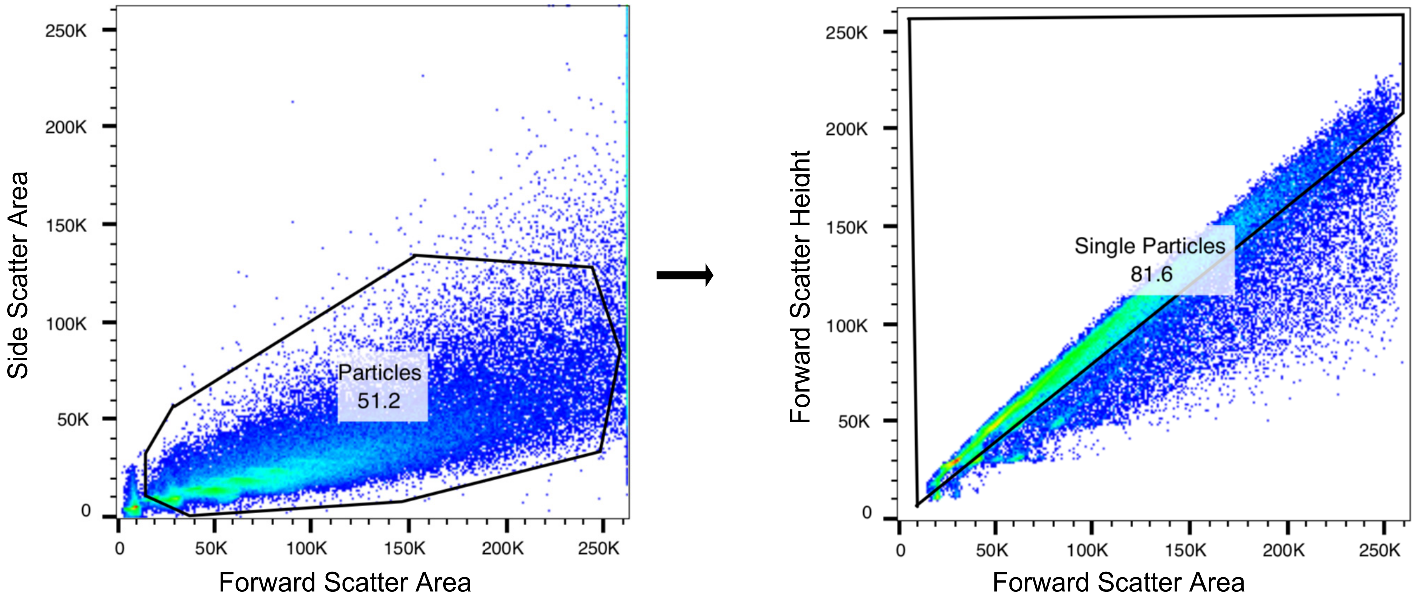


**Supplementary Figure 7. Gating Strategy for flow cytometry for fluorospheres.** Gating strategy on the analysis of *in vitro* binding assay (**Fig. 5**).
